# Negative Impact of Cenobamate on Cognition: Dose-Dependent and Independent Effects

**DOI:** 10.1101/2024.12.23.24319533

**Authors:** Juri-Alexander Witt, Mostafa Badr, Rainer Surges, Randi von Wrede, Christoph Helmstaedter

## Abstract

**Background:** Studies on Cenobamate (CNB) have generally reported neutral to positive effects on objective cognitive performance in patients with epilepsy, but are limited to dosages up to 250 mg/day. However, a case report (Witt et al., *Neurocase*, 2024) noted severe memory deterioration at 400 mg/day.

**Objective:** To examine dose-dependent effects of CNB on cognition.

**Methods:** In this retrospective longitudinal study, executive functions (EpiTrack®) and episodic memory were assessed in patients with epilepsy during CNB therapy and compared to baseline. Subgroups were stratified by daily CNB doses of ≥300 mg versus <300 mg.

**Results:** The study included 84 patients. With a mean CNB dose 200.6 ± 114.3 mg (range: 12.5-400.0 mg; 28.6% ≥300 mg) seizure freedom was achieved in 10.7%. Repeated measures ANCOVA revealed a significant decline in executive functions at ≥300 mg (n=84; F = 6.35, p = 0.014). Changes were correlated with CNB dose (r = −0.31, p = 0.004). Significant individual declines according to reliable change indices occurred in 50.0% of patients on higher versus 16.7% on lower CNB doses. In a subgroup undergoing extensive memory testing, verbal retention showed a significant negative, dose-independent effect (n=22; F = 7.95, p = 0.011), with intraindividual declines in 28.6% (≥300 mg) versus 13.3% (<300 mg). Other memory parameters were unaffected.

**Conclusion:** In the largest longitudinal study to date, higher CNB doses were linked to deterioration in executive functions, while a decline in verbal retention appeared dose-independent. These findings need to be confirmed by systematic studies.

## 1 Introduction

Antiseizure medications (ASM) can positively impact cognitive function primarily through effective seizure control and reduction of interictal epileptic discharges [1]. However, cognitive side effects that may impair daily functioning, hinder long-term retention, and reduce quality of life are more common [2,3]. Cognitive and psychiatric side effects are among the least tolerated adverse effects of ASMs [4], and the likelihood of these side effects varies across different ASMs and increases with polytherapy and higher drug loads [1,5–8]. Additionally, idiosyncratic reactions may also occur. The cognitive domain of attention and executive functions is typically the most vulnerable and therefore also the most sensitive in relation to ASM-induced side effects [1].

Cenobamate (CNB) is a newer ASM approved for treating focal-onset seizures. Its mechanism of action include inhibition of voltage-gated sodium channels and activation of extrasynaptic GABA-receptors [9], showing promising efficacy in patients with refractory epilepsy [10,11]. Common treatment-emergent adverse effects include somnolence, dizziness, fatigue, and diplopia [12]. A pooled analysis from phase 2 and phase 3 clinical trials identified cognitive adverse events in up to 1.9% of patients on adjunctive CNB, compared to 0.5% with placebo with disturbances in memory, attention, and language [13].

Up to now, there are three group studies in patients with epilepsy and one case report on the cognitive effects of CNB using standardized tests. In a prospective observational study from Germany [14], cognitive assessments of 50 patients with pharmacoresistant focal epilepsy were conducted before and three months after starting adjunctive CNB (50–250 mg/d). The assessment employed the EpiTrack®, a screening tool devised to monitor cognitive effects of ASM, as sole measure. Results indicated group-level improvements in attention and executive function, with stable performance for most individuals (72%). Significant cognitive declines occurred in 12% of the patients, while 16% showed significant improvement. Changes in cognition were not correlated with CNB dose, cumulative drug load, or seizure control (seizure freedom achieved in 6% of the patients).

In a prospective observational study from Spain by Catalán-Aguilar and colleagues [15], 32 patients with pharmacoresistant focal epilepsy were cognitively tested before and 3 months (T1) after CNB administration (50–200 mg/d). A subsample of 22 patients was reassessed again after 6 months (150–250 mg/d). The cognitive evaluation included tests on attention and executive functions (EpiTrack®), memory (Wechsler Memory Scale III, WMS-III), and language (Boston Naming Test, BNT). Group analyses did not indicate significant changes under CNB. Seizure freedom (28.0% after 3 months, 13.6% after 6 months) was not associated with changes in cognition.

An explorative study from another group from Spain [16] retrospectively analyzed cognition in 20 patients with pharmacoresistant focal epilepsy before and after reaching the first target dose of CNB after 6 months. Mean CNB doses were 12.5 mg/day at baseline and 191.2 mg/day at 6 months. Analyses on a group level showed statistically significant improvement in two measures of verbal episodic and one measure of visuospatial episodic memory, whereas they also report a significant worsening in visuo-motor speed.

In contrast to this, a recent case report from our department [17] describes a patient who experienced a dose-dependent, severe decline in episodic memory under 400 mg/day of CNB, leading to a collapse in school performance, while his attention and executive functions remained stable and unaffected. After halving the dose, objective memory performance fully recovered. An animal study [18] previously reported a negative impact of CNB on novel object recognition, object location memory, and spatial learning in mice.

Against this background our retrospective longitudinal study investigates potential dose-dependent effects of CNB on objective cognitive performance. We hypothesize a negative impact particularly on episodic memory performance at higher CNB doses.

## 2 Methods

### 2.1 Study design and patients

In this retrospective longitudinal study objective cognitive test performance of patients with pharmacoresistant epilepsy were assessed during CNB therapy and compared to baseline. To reveal potential dose-dependent effects, subgroups were stratified by daily CNB doses of ≥300 mg versus <300 mg.

We searched our local ASM registry (approval of the Ethics commission of the Medical Faculty at the University of Bonn (No. 130/19)) for all patients who underwent a cognitive assessment under CNB. Inclusion criteria included a diagnosis of epilepsy according to ILAE guidelines, available cognitive baseline data without CNB, available information on concomitant ASM. Patients were excluded when brain damage occurred or invasive interventions (e.g. stereo EEG) or epilepsy surgery were performed in the time between the assessment with and without CNB or in case of known progressive pathologies. When a patient was tested under different CNB doses, the assessment with the highest dose was chosen.

### 2.2 Cognitive assessment

The cognitive examinations focused on executive functions and episodic memory. Executive functions and attention were assessed via the EpiTrack® (third edition) [19], a screening tool particularly sensitive to medication effects, making it highly suitable for monitoring of pharmacological treatments [20,21]. It comprises six subtests evaluating response inhibition, visuo-motor speed, mental flexibility, visual-motor planning, verbal fluency, and working memory. An age-adjusted total score is calculated based on subtest results. Scores between 29 and 31 indicate mild impairment, while significant impairment is defined as a score of ≤28. According to reliable change indices (RCIs) significant change in performance is reflected by an improvement of >3 points or a decline of >2 points. Research supports the utility of EpiTrack® for cognitive monitoring of pharmacological treatments [22–28] and its sensitivity to overall drug load, including the number of concurrent ASM [7,8].

Episodic memory in the verbal domain was assessed using the Verbaler Lern-und Merkfähigkeitstest (VLMT) [29], a modified German version of the Rey Auditory Verbal Learning Test – RAVLT [30] or via an abbreviated version with two learning trials before and delayed free recall after the EpiTrack® [24–26]. The VLMT is the most frequently applied verbal learning and memory test in German epilepsy centers [31]. Analyses were based on learning performance (total number of words learned over all learning trials), and memory retention (loss of words over the retention interval). In addition, the standard VLMT evaluates recognition performance (corrected for false-positive answers). VLMT raw scores were transformed into age-corrected standard values (mean = 100.0, standard deviation = 10.0) according to normative data. The raw scores of the abbreviated VLMT were converted into a scale ranging from 1 to 7 according to the normative data and merged into a total memory score [24]. Regarding the standard VLMT, parallel forms were employed at the reassessment. Practice-corrected RCIs were used to determine statistically significant intraindividual changes. The VLMT has been shown to be sensitive to left temporal lobe dysfunction, left mesiotemporal pathology and left-sided temporal lobe surgery [32–35].

Nonverbal episodic memory was evaluated using the revised version of the *Diagnosticum für Cerebralschädigung* (DCS-R) [36]. This test involves learning and reconstructing nine abstract designs over five consecutive trials, followed by a recognition memory assessment after a 30-minute retention interval. Analyses were based on the number of correctly learned designs across the five learning trials and recognition performance, adjusted for false-positive responses. Analogue to the VLMT, raw scores were transformed into age-corrected standard values (mean = 100.0, standard deviation = 10.0). Parallel forms were employed at the reassessment. Practice-corrected RCIs were applied to evaluate statistically significant intraindividual changes. The DCS-R has demonstrated sensitivity to dysfunctions of the right temporal lobe, right mesiotemporal pathology, and right-sided temporal lobe surgery [33,36–38].

### 2.3 Statistical analyses

Data were analyzed using IBM SPSS Version 29 and JASP 0.18.3.0. Descriptive statistics were calculated to summarize demographic and clinical characteristics, including frequencies, means and standard deviations. Repeated measures analysis of covariance (ANCOVA) was employed to evaluate within-subject changes in cognitive performance under CNB treatment, with CNB dose (≥300 mg versus <300 mg) included as a between-subject factor. Covariates, such as changes in the number of concurrent ASMs, were controlled to account for potential confounding effects. Pearson correlation coefficients were calculated to examine the relationship between CNB dose and changes in cognitive performance. Reliable change indices (RCIs) were applied to evaluate significant intraindividual changes in cognitive test scores. Further statistics included paired t-tests and chi²-tests. Statistical significance was set at p < 0.05 for all tests.

## 3 Results

After exclusion of patients with intermediate brain damage (n = 2), after treatment with radiofrequency-thermocoagulation (n = 3), deep brain stimulation (n = 1), or invasive EEG monitoring (n = 5) and those with a progressive pathology (n = 2), lack of a baseline assessment without CNB (n = 23) or incomplete or incomparable cognitive assessments (n = 3), the final sample consisted of 84 patients with epilepsy. Major demographic and clinical characteristics of the sample are presented in **Table 1**.

**Table 1:** Demographic and clinical characteristics.

|  | Total | CNB < 300 mg/d | CNB ≥ 300 mg/d |  |
| --- | --- | --- | --- | --- |
| N | 84 | 60 | 24 |  |
| Sex |  |  |  |  |
| Female | 47 (56.0%) | 34 (56.7%) | 13 (54.2%) | Chi<sup>2</sup> = 0.04,<br>p = 0.835 |
| Male | 37 (44.0%) | 26 (43.3%) | 11 (45.8%) |  |
| Age |  |  |  |  |
| M (SD) yrs | 39.7 (13.0) | 40.6 (12.9) | 37.6 (13.5) | T = 0.96,<br>p = 0.341 |
| Range | 18-71 | 21-71 | 18-66 |  |
| Age at onset of epilepsy |  |  |  |  |
| M (SD) yrs | 16.3 (11.9) | 15.1 (10.1) | 19.4 (15.3) | T = -1.26,<br>p = 0.215 |
| Range | 0-62 | 0-47 | 1-62 |  |
| Epilepsy diagnosis |  |  |  |  |
| Structural | 60 (71.4%) | 46 (76.7%) | 14 (58.3%) | Chi<sup>2</sup> = 3.71,<br>p = 0.156 |
| Unknown | 23 (27.4%) | 13 (21.7%) | 10 (41.7%) |  |
| Generalized genetic | 1 (1.2%) | 1 (1.7%) | 0 (0.0%) |  |
| Number of ASMs at baseline |  |  |  |  |
| M (SD) | 2.7 (1.0) | 2.8 (0.9) | 2.6 (1.1) | T = 0.528,<br>p = 0.599 |
| Range | 1-5 | 1-5 | 1-5 |  |
| Number of ASMs under CNB |  |  |  |  |
| M (SD) | 3.2 (0.9) | 3.2 (0.9) | 3.1 (0.9) | T = 0.50,<br>p = 0.622 |
| Range | 2-5 | 2-5 | 2-5 |  |
| Defined daily dose (DDD) at baseline |  |  |  |  |
| M (SD) | 3.8 (1.8) | 3.7 (1.7) | 3.9 (2.1) | T = -0.42,<br>p = 0.673 |
| Range | 1-11 | 1-11 | 1-9 |  |
| Defined daily dose (DDD) under CNB |  |  |  |  |
| M (SD) | 4.3 (1.9) | 4.0 (1.7) | 4.4 (2.3) | T = -2.45,<br>**p = 0.016** |
| Range | 2-13 | 2-10 | 2-13 |  |
| CNB dose (mg/d) |  |  |  |  |
| M (SD) | 200.6 (114.3) | 142.5 (73.2) | 345.8 (51.0) | T = -12.44,<br>**p < 0.001** |
| Range | 12.5-400 | 12.5-250 | 300-400 |  |

CNB, cenobamate; ASMs, antiseizure medications; DDD, defined daily dose; M, mean; SD, standard deviation

The mean CNB daily dose was 200.6 ± 114.3 mg (range: 12.5-400.0 mg) and 24 patients (28.6%) had a high CNB dose ranging between 300 and 400 mg. Seizure freedom under CNB was achieved in 10.7% of the patients and there was no statistical significant difference between the low and the high dose subgroup (high: 8.3% vs. low: 11.7%, Chi² = 0.20, p = 0.655). The number of combined ASMs was higher during the assessment with CNB as compared to baseline (3.2 versus 2.7 ASMs, T = −5.82, p < 0.001; **Table 2**). Therefore, this difference was included as a covariate in the analyses. Changes in co-medication are listed in **Table 2**. Along with the introduction of CNB (+100%), an increase of brivaracetam (BRV; +11.9%) use and a decrease of levetiracetam (LEV; −15.5%), oxcarbazepine (OXC; −8.3%) and perampanel (PER; −6.0%) prescriptions were observed.

**Table 2:** Antiseizure medication (ASM) at baseline and under cenobamate (CNB) therapy.

|  | Baseline | Cenobamate |  |
| --- | --- | --- | --- |
| N | 84 | 84 |  |
| Number of ASMs at |  |  |  |
| M (SD) | 2.7 (1.0) | 3.2 (0.9) | T = -5.82, |
| Range | 1-5 | 2-5 | **p < 0.001** |
| Defined daily dose (DDD) |  |  |  |
| M (SD) | 3.8 (1.8) | 4.3 (1.9) | T = -3.19, |
| Range | 1-11 | 2-13 | **p = 0.002** |
| Concomitant ASMs |  |  |  |
| Brivaracetam (BRV) | 28 (33.3%) | 38 (45.2%) | ↑↑ |
| Cannabidiol (CBD) | 9 (10.7%) | 7 (8.3%) | → |
| Carbamazepine (CBZ) | 6 (7.1 %) | 3 (3.6%) | → |
| Cenobamate (CNB) | 0 (0.0%) | 84 (100.0%) | ↑↑ |
| Clobazam (CLB) | 14 (16.7%) | 15 (17.9%) | → |
| Clonazepam (CLZ) | 4 (4.8%) | 2 (2.4%) | → |
| Diazepam | 0 (0.0%) | 1 (1.2%) | → |
| Eslicarbazepin acetate (ESL) | 12 (14.3%) | 8 (9.5%) | → |
| Lacosamide (LCM) | 18 (21.4%) | 14 (16.7%) | → |
| Lamotrigine (LTG) | 36 (42.9%) | 33 (39.3%) | → |
| Levetiracetam (LEV) | 31 (36.9%) | 18 (21.4%) | ↓↓ |
| Lorazepam (LZP) | 3 (3.6%) | 0 (0.0%) | → |
| Oxcarbazepine (OXC) | 10 (11.9%) | 3 (3.6%) | ↓ |
| Perampanel (PER) | 22 (26.2%) | 17 (20.2%) | ↓ |
| Phenobarbital (PB) | 4 (4.8%) | 3 (3.6%) | → |
| Pregabalin (PGB) | 1 (1.2%) | 1 (1.2%) | → |
| Phenytoin (PHT) | 4 (4.8%) | 2 (2.4%) | → |
| Retigabine (RTG) | 1 (1.2%) | 0 (0.0%) | → |
| Sultiame (STM) | 1 (1.2%) | 0 (0.0%) | → |
| Topiramate (TPM) | 4 (4.8%) | 2 (2.4%) | → |
| Valproic acid (VPA) | 9 (10.7%) | 10 (11.9%) | → |
| Zonisamide (ZNS) | 11 (13.1%) | 8 (9.5%) | → |

ASMs, antiseizure medications; DDD, defined daily dose; M, mean; SD, standard deviation; ↑↑, increase > 10%; ↑, increase > 5% →, changes < 10%; ↓, decrease > 5%; ↓↓, decrease > 10%

Repeated measures ANCOVA indicated a significant decrease in executive performance (EpiTrack®) in those with ≥300 mg of CNB (interaction effect: F = 6.3, p = 0.014; **Figure 1**). Performance changes in EpiTrack® correlated with CNB dose (r = −0.31, p = 0.004). At the individual level, significant declines (RCI) in performance were observed in 50.0% of patients on higher CNB doses (compared to 16.7% on <300 mg/day; improvements: 4.2% versus 13.3%; Chi² = 10.20, p = 0.006; **Figure 2**). In the memory domain, only verbal retention showed a significant negative effect, as assessed by the VLMT (n = 22; F = 7.9, p = 0.011; **Figure 3**) and found to be independent of CNB dose. The incidence of significant individual declines (RCI) was 28.6% for higher doses and 13.3% for lower doses (Chi² = 0.75, p = 0.388), with no significant individual improvement observed. Other memory parameters and tests remained unaffected by CNB (**Table 3**).

**Figure 1:**
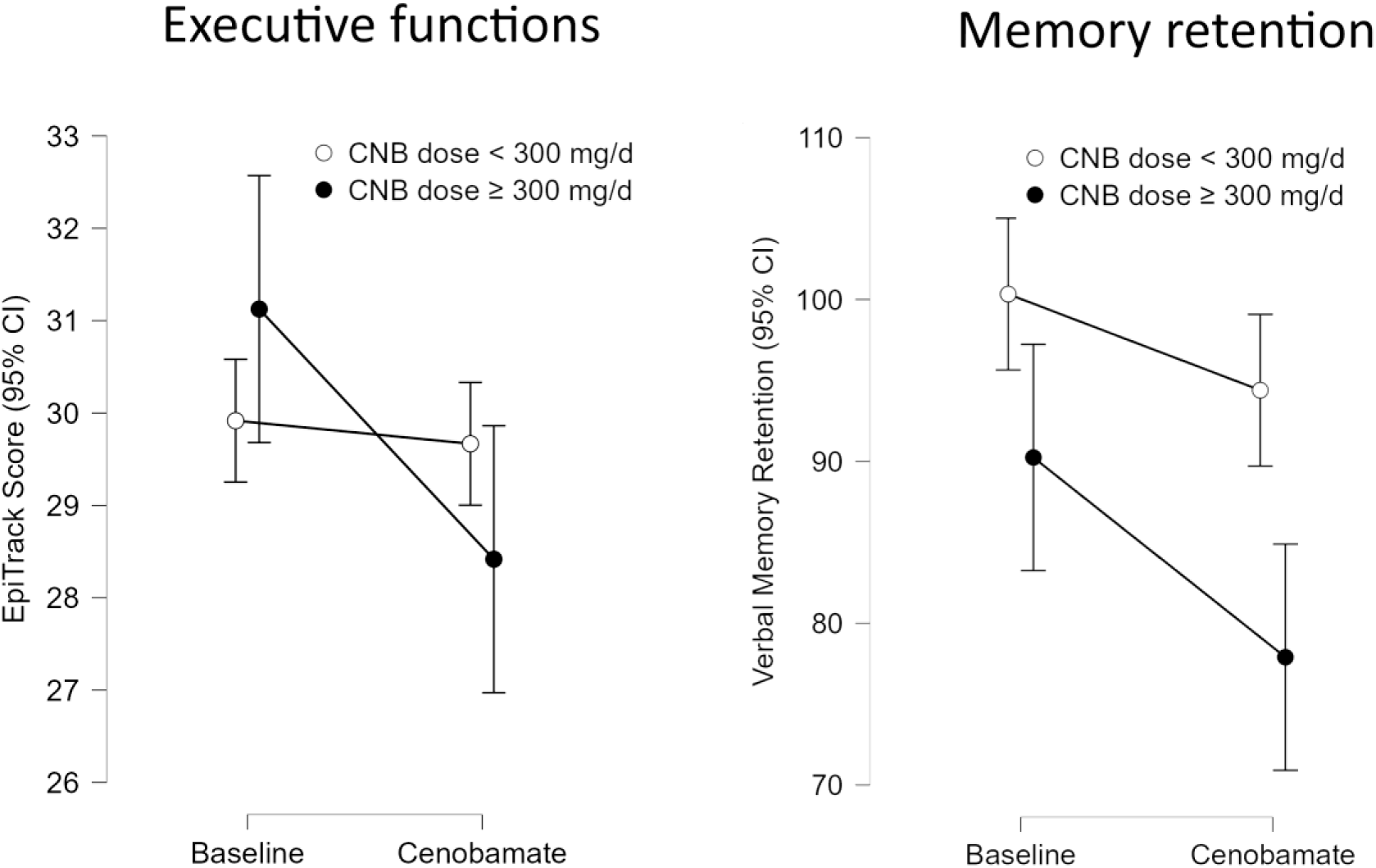
Change in executive functions (EpiTrack®) and memory retention (VLMT) during cenobamate (CNB) therapy as compared to baseline. The figure on the left shows a significant interaction effect (n=84; F = 6.3, p = 0.014) in terms of a selective decline in executive functions in patients with a high CNB dose (≥ 300 mg/d). The figure on the right depicts a dose-independent decline in memory retention (VLMT) in a subgroup of patients who underwent an extensive memory assessment (n=22; F = 7.9, p = 0.011). Depicted are the estimated means and 95% confidence intervals (CI) after statistical control for difference in the number of antiseizure medications (ASM) between baseline and assessment during CNB therapy.

**Figure 2:**
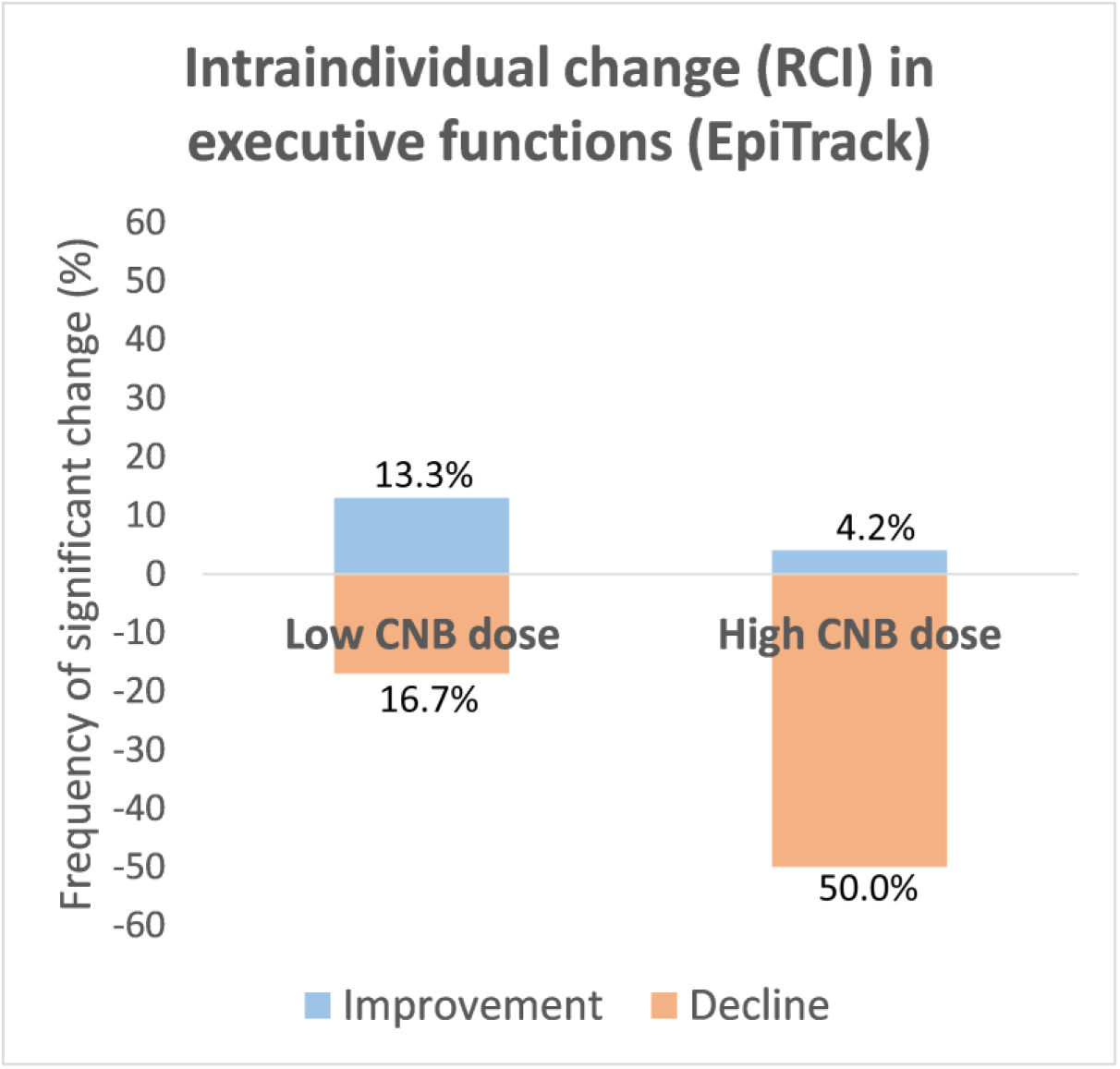
Percental frequencies of significant intraindividual change in executive functions (EpiTrack®) under cenobamate (CNB) according to reliable change indices (RCI). An intraindividual decline was more frequently and improvement less often observed in the subgroup of patients with high CNB doses (≥ 300 mg/d) (Chi² = 10.20, p = 0.006).

**Figure 3:**
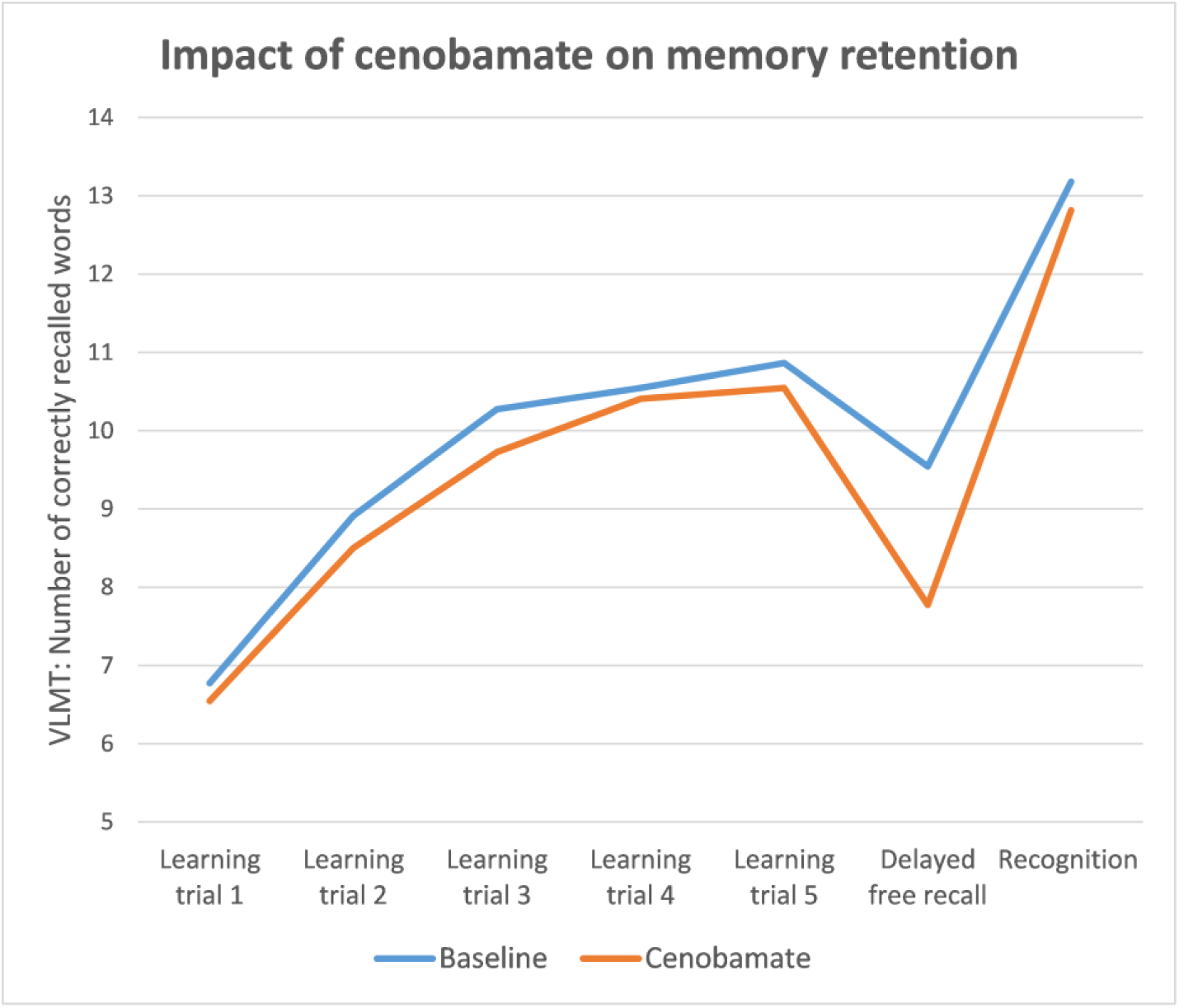
Change in verbal learning and memory performance under cenobamate (CNB). Analyses indicated a significant selective negative effect on memory retention (n = 22; F = 7.9, p = 0.011) which was not dose-dependent.

**Table 3:** Cognitive performance at baseline and during therapy with cenobamate (CNB) as well as frequencies of significant intraindividual changes according to reliable change indices (RCIs).

|  | N | Baseline | Cenobamate | Statistics |  | Significant intraindividual changes (RCI) |  |
| --- | --- | --- | --- | --- | --- | --- | --- |
|  |  | M (SD) | M (SD) | Main effect<sup>a</sup> | Interaction effect (CNB dose group)<sup>b</sup> | ↓ | ↑ |
| Executive functions<br>EpiTrack | 84 | 30.3 (5.4) | 29.3 (6.8) | F = 5.88, **p = 0.018** | F = 6.35, **p = 0.014** | 26.2% | 10.7% |
| Episodic memory |  |  |  |  |  |  |  |
| VLMT |  |  |  |  |  |  |  |
| Learning | 22 | 89.2 (16.2) | 89.6 (15.4) | F = 1.13, p = 0.301 | F = 0.60, p = 0.447 | 31.8% | 13.6% |
| Retention | 22 | 97.1 (15.9) | 89.1 (14.5) | F = 7.95, **p = 0.011** | F = 1.41; p = 0.250 | 18.2% | 0.0% |
| Recognition | 22 | 81.7 (25.8) | 78.3 (28.8) | F = 0.03, p = 0.870 | F = 0.29; p = 0.598 | 36.4% | 18.2% |
| VLMT (abbreviated)<br>....Total score | 34 | 13.1 (2.5) | 13.7 (2.5) | F = 0.06, P = 0.816 | F = 1.48, p = 0.233 | 0.0% | 8.8% |
| DCS-R |  |  |  |  |  |  |  |
| Learning | 21 | 82.9 (15.4) | 82.5 (14.0) | F = 1.34, p = 0.263 | F = 0.24, p = 0.628 | 14.3% | 9.5% |
| Recognition | 20 | 89.0 (18.1) | 92.5 (19.0) | F = 0.29, p = 0.596 | F = 0.11, p = 0.734 | 0.0% | 15.0% |

CNB, cenobamate; M, mean; SD, standard deviation; ↓, decline; ↑, improvement; <sup>a</sup>, repeated measures analysis of covariance (ANCOVA) accounting for intraindividual changes in the number of concurrent ASMs as covariate; <sup>b</sup>, interaction effect with low versus high dose of CNB

## 4 Discussion

The study contributes to the growing body of literature on the cognitive effects of CNB, a novel ASM approved as adjunctive treatment for focal-onset seizures. Our findings demonstrate a dose-dependent negative impact of CNB on attentional-executive functions, as measured by the EpiTrack®, whereas episodic memory performance in terms of verbal retention showed significant but dose-independent declines. On the other hand, seizure freedom was achieved in 10.7% of the patients without a significant effect of CNB dose. These results point to potential cognitive trade-offs associated with an antiseizure treatment with CNB, which must be carefully weighed against its efficacy in seizure control.

### 4.1 Impact of CNB on executive functions

The observed dose-dependent decline in executive performance was unexpected given the findings from previous studies which indicated a neutral or even positive effect of CNB on executive functions at a group level. However, these studies were limited to dosages up to 250 mg/day, but according to our study deteriorations in executive functions seem to occur with rather higher doses. Notably, 50.0% of patients on higher CNB doses exhibited significant declines in this domain, a rate substantially higher than in the low-dose group (16.7%). The latter was comparable with the results of the studies by Schuetz and colleagues [14] and by Catalán-Aguilar et al. [15] with 12.0-12.5% of patients showing a significant decline at the 3-month follow-up. Thus, when considering the CNB dosage, there is actually no discrepancy between our and the previous studies. Moreover, in the study by Catalán-Aguilar and colleagues [15] at least 25% of the patients significantly deteriorated in the Trail Making Test A (TMT-A), a subtest of the EpiTrack® and a measure of visuo-motor speed. Notably, Serrano-Castro et al. [16] found a slight but significant decline under CNB in the very same test at a group level. We, however, reported a case with a marked decline in episodic memory under 400 mg/day of CNB, leading to a collapse in school performance, while performance in executive functions remained stable and unaffected [17].

### 4.1 Impact of CNB on episodic memory

Our current findings may suggest that CNB also selectively affects verbal retention without significantly impairing other aspects of episodic memory, such as learning or recognition. This was seen in the subgroup of patients undergoing extensive memory testing. Up to now only two previous group studies also assessed episodic memory before and after introduction of CNB. Catalán-Aguilar and colleagues [15] assessed verbal memory using the WMS-III and did not find any deteriorations under CNB, neither at a group nor at an individual level. However, the WMS-III does not provide alternative forms of the memory contents and thus the very same memory items were presented at baseline and the (up to) two follow-ups. It cannot be excluded that a learning effect may have masked potential negative effects of CNB. The WMS-III is therefore not suited for longitudinal studies [39,40]. Moreover, performance in the WMS seems to depend more on non-memory domains such as executive functions and semantic memory and less sensitive to hippocampal dysfunction than the VLMT [41]. Serrano-Castro et al. [16] employed the Free and Cued Selective Reminding Test (FCSRT) to assess verbal episodic memory and the Rey-Osterrieth Complex Figure Test (ROCFT) to evaluate visuospatial episodic memory. Parallel forms were used to minimize learning effects and both tests showed a significant improvement with medium effect sizes at a group level. There are clear differences between the FCSRT and the VLMT, e.g. in the FCSRT only 4 words need to be learned in three learning trials, whereas the VLMT requires learning of 15 words in 5 trials. Thus the VLMT appears more demanding and its use in the field of epilepsy is strongly evidence-based (see section 2.2). Finally, a mouse study [18] indeed reported negative effects of CNB on novel object recognition, object location memory, and spatial learning as compared to controls. More systematic studies in patients with epilepsy using appropriate measures are needed to confirm negative effects of CNB on episodic memory.

### 4.4 Limitations

Limitations include the retrospective study design, limited generalizability given the treatment resistance of patients from a specialized epilepsy center, the sometimes large time gap between testing sessions (patients with relevant medical events or invasive treatments in-between or progressive pathologies were excluded), additional but mostly limited changes in co-medication (**Table 2**; analyses considered differences in numbers of concurrent ASMs as covariate), and the use of different versions of the verbal memory test (leading to subsample analyses with lower statistical power).

## 5 Conclusion

This study demonstrates a dose-dependent negative impact of CNB on executive functions and preliminarily points to potential dose-independent adverse effects on memory retention. Our findings underscore the importance of recognizing and addressing the cognitive side effects of CNB, particularly at higher doses. While the medication offers promising efficacy for seizure control, its cognitive impact necessitates careful dose titration. We recommend to prioritize the lowest effective dose of CNB, balancing seizure control against potential cognitive side effects, and consider routine cognitive monitoring.

## Data Availability

The dataset analyzed during the current study is not publicly available.

## Author contributions

JAW conceptualized the study. MB, RS and RvW, were involved in the acquisition of patients and provided clinical data. JAW performed the data analysis and prepared the first draft of the manuscript. All authors revised the manuscript for intellectual content, approved the final version of the manuscript and are accountable for their work.

